# Prenatal Exposure to Compliant Nitrate in Iowa Public Drinking Water (1983-1988)

**DOI:** 10.64898/2026.09.14.26363048

**Authors:** Jason Semprini

**Author notes:** **Correspondence:** Jason Semprini, 8025 Grand Avenue, West Des Moines, IA 50266.

## Abstract

Essential for nutrient growth, due to intensive farming practices nitrate has become a common water contaminant in agricultural states. US nitrate regulations are designed to protect the health of newborns and infants, but have never been revised to consider potential harms of prenatal exposure. Recent evidence, however, suggests that exposure below the 10 mg/L regulatory limit may be associated with adverse birth outcomes. This current study examines whether first-trimester exposure below the regulatory limit is associated with adverse birth outcomes in Iowa (an intensive agricultural state). Finished drinking-water nitrate measurements from Iowa public water systems were linked to birth records. Nitrate measures were aggregated to a balanced county-quarter panel (1983 to 1988). Each pregnancy was assigned an average nitrate exposure over a 98-day first-trimester window. All births with a first trimester county-quarter nitrate measure >= 10 mg/L were excluded. Linear probability models with county-by-year and year-by-conception-quarter fixed effects, with standard errors clustered by county, estimated both uncorrected and Sidak corrected inference. The final analytic sample included 113,328 singleton births. Relative to lowest exposure category (0.1 mg/L or below), exposure to 5 mg/L or more nitrate was associated with a 1.04 percentage-point increase in the probability of preterm birth (CI = 0.24 to 1.83), a 13.5% relative increase, and associated with a 0.53 percentage-point increase in very preterm birth (CI = 0.08 to 0.97), a 46.2% relative increase. The statistical significance of both estimates persisted after correction for multiple tests. Exposure to > 0.1 mg/L and < 5 mg/L was not associated with any outcome. These preterm and very preterm birth associations were consistent across several sensitivity checks. Because nitrate concentrations between 5 to <10 mg/L comply with current rules, these findings warrant reassessing approaches to protect the health of pregnant women and their children.

## Background

Naturally occurring, but widely used in synthetic nitrogen fertilizer, nitrate remains an essential nutrient for crop growth[1,2]. However, with greater nitrogen application, nitrates can accumulate in the soil and then seep into groundwater[3]. In agricultural regions, nitrate has become a widespread chemical contaminant in water risking possible harms to human health[4–6]. To reduce impact on health, the US federal government has set a maximum contaminant level (MCL) of nitrates in drinking water at 10 mg/L[7]. The US Environmental Protection Agency (EPA) explicitly states that infants drinking water with elevated nitrates can lead to methemoglobinemia (Blue Baby Syndrome)[7,8]. The EPA does not mention any risk below the MCL or for any other population at risk of drinking water with elevated nitrates. Yet, evidence continues to emerge that prenatal exposure to nitrates may be associated with increased risk of adverse birth outcomes, even exposure to compliant levels of nitrate[9–11].

Nitrate, by converting to nitrite, presents highly plausible, biological harm[8]. Such possible harms have been assessed by numerous studies in humans, revealing inconsistent and mixed evidence as to the effect of consuming nitrates in drinking water[6,9,12]. This inconsistency likely stems from inherent challenges to evaluating the effect of non-random exposure in humans with observational data and non-causal designs. Two recent studies aimed to overcome such limitations with causal inference econometric methods, both finding that exposure to nitrate levels below the 10 mg/L MCL threshold were associated with increased risk of preterm birth and low birthweight[10,11]. Both of these studies were set in California, which may not adequately represent the burden of exposure to compliant nitrates in public drinking water across other agricultural regions.

The intensively farmed states of the Midwest differ from California on important dimensions. For one, California’s drinking water infrastructure is quite unique in its diversity[13]. Further, Midwest application rates of nitrogen fertilizer are among the highest in the country and human exposure in drinking water varies seasonally[5,14,15]. In addition to intense, seasonal nitrogen fertilizer application, Midwest agricultural states have increased nitrate exposure due to greater prevalence of animal feeding operations[16]. Still, likely due to differences in settings and methods, population-based evidence on prenatal nitrate exposure in these high-agriculture settings remains minimal[17–20].

### Objective

In 2025, Semprini published a population-based, individual-level study in a high-agriculture state, linking Iowa birth records from 1970–1988 to public water system nitrate monitoring data and implementing a causal inference design[21]. However, in 2026, Semprini’s study was retracted as the analysis included both finished and unfinished drinking water measures[22]. A number of other issues related to exposure, compliance, and correcting for multiple tests were also identified in the retraction[22]. The following study aims to update Semprini’s 2025 study by addressing these key issues, transparently correcting the scientific record, and presenting evidence of nitrate’s potential harms to pregnant women and children in a state with some of the highest nitrate levels in the US[23–25].

### Updates to 2025 Study

#### Finished Water Samples Only

The stated reason for retraction was that the water data used to construct the exposure variable included unfinished water, raw and compliance check measures, alongside finished-water samples. This distinction is consequential, as raw nitrate measures may not reflect nitrate ultimately found in public drinking water systems[26]. Nitrate concentrations in raw water samples could be reduced with treatment, so a measure of both finished and unfinished water samples may not reflect true nitrate exposure. Correcting this error was the primary motivation for the present study, which was restricted to public drinking water measures classified as finished.

#### Measurements at or above the regulatory limit

The original study also retained nitrate measures at or above 10 mg/L. This decision ignored possible reporting and remediation, which could not be measured with the historic data. Again, the nitrate measures may not reflect public drinking water delivered to or consumed by the public. After linking nitrate measures to birth records, this present study excluded all birth records exposed to non-compliant nitrate measures above the 10 mg/L threshold.

#### 30-day exposure window

Semprini (2025) defined exposure to nitrate over a 30-day window of estimated conception. This approach also may not accurately represent true exposure. Following the examples of existing research, the present study defines exposure to nitrate over a fixed first-trimester window spanning gestational days 0–97 (98 days).

#### Multiple hypothesis testing

Finally, Semprini (2025) reported unadjusted confidence intervals across multiple exposure categories without correction for multiplicity. Although not commonly performed in the existing evidence base[10,11], the present analysis reports the unadjusted quantities, following the reporting practice of Sherris et al. (2021) and Montoya (2025), but also reports adjusted confidence intervals and p-values using the Šidák correction[27].

## Methods

### Data

Drinking-water nitrate measurements were obtained from the Center for Health Effects of Environmental Contamination (https://cheec.uiowa.edu/data), the same source used in Semprini (2025). This archived data compiles nitrate analyses submitted by Iowa public water systems, and includes the system identifier, the sampling date, the measured nitrate concentration, and the geographic coordinates of the sampling location. Only finished samples were included in the present analysis.

See data availability statement for information on accessing the original data source of finished water samples, which spanned 1970-1988. However, water sample data was notably sparse and inconsistent prior to 1983. Therefore, the final water data was restricted to 1983-1988.

Birth records (1983-1988) were drawn from the National Center for Health Statistics natality detail files distributed by the National Bureau of Economic Research (https://www.nber.org/research/data/vital-statistics-natality-birth-data), covering all births occurring in Iowa. These files provide the date of birth, county of maternal residence, gestational age, birthweight, plurality, infant sex, and maternal age, parity, education, race, marital status, and month of prenatal care initiation.

### Construction of Quarterly-County Panel of Nitrate Measures

The water data was restricted to finished-water samples. Nitrate was measured as N mg/L as nitrogen. Nitrate values at or above 10 mg/L were initially retained without deletion or truncation at every stage of panel construction, but all county-quarters with an observed nitrate measure at or above 10 mg/L were flagged for exclusion after linkage to birth records. Using the water measure sampling coordinates, each water system was assigned to its Iowa county. To prevent water systems that sample frequently or allow extreme outlier measures to dominate county-level exposure, measurements were aggregated in a fixed hierarchy rather than averaged directly. Raw analyses were summarized 1) median within water system-sampling-day, 2) median water system-month, 3) median water system-quarter, and 4) median across systems to the county-quarter. In the end, a balanced panel of all 99 Iowa counties by all calendar quarters from 1983-Q1 through 1988-Q4 (2,376 county-quarters) was constructed.

Between 1983-1988, nitrate monitoring was intermittent. 30.7% of all county-quarters had no nitrate measure. Following a similar method to Sorensen Montoya[11], the panel was completed using a spatiotemporal regression model on the log-transformed scale. The specification included county fixed effects to account for time-invariant spatial differences in nitrate concentrations, year-quarter fixed effects to capture common temporal variation, and the log-transformed contemporaneous mean nitrate concentration in neighboring counties as a spatial predictor. The spatial predictor for a given county-quarter is the inverse-distance-weighted mean of observed concentrations in other counties in the same quarter within 100 km, excluding the target county itself; where no observed county lay within 100 km, the five nearest observed counties in that quarter were used. Predictions were returned to the natural scale using a Duan smearing factor computed from the model residuals, an appropriate approach for predictive environmental modelling[28,29].

The predictive model uses water monitoring data, geography, and calendar time. Birth records, maternal characteristics, and birth outcomes never enter the model. So, any error introduced by the prediction model is non-differential with respect to the birth outcomes and is consistent with spatiotemporal approaches used to address irregularly sampled environmental monitoring data[30]. Further, as described below, birth records without at least one county-quarter nitrate measure during the first trimester were excluded. Observed values were never replaced by predicted values. The resulting panel yielded the observed concentration where one exists and the completed prediction otherwise.

The completed prediction model was validated under three holdout designs. In every replication, the spatial predictor was rebuilt from training counties and the Duan smearing factor was recomputed from training residuals. The first design was a random five-fold cross-validation. The second design held out a randomly chosen pair of contiguous quarters within each county. The third design imposed artificial missingness on observed counties, retaining a random 75%, 50%, or 25% of each county’s observed quarters and repeating each retention rate 30 times.

Sensitivity (see below) is eventually assessed using an exposure measure constructed from directly observed county-quarters.

72.0% of the pregnancies in the final analytic data had observed county-quarter nitrate measures spanning all 98 days of the first trimester. 28% of the pregnancies required the completed county-nitrate measures to assemble the full first trimester mean exposure. The observed and completed county-quarter nitrate means were 2.7 and 2.8 mg/L, respectively, with a mean difference of 0.1 mg/L between observed and completed nitrate measures.

### Creating and Linking Birth Records

Exact birth dates were reconstructed from recorded year, month, and day. Gestational age was assigned by a stated hierarchy: where an exact last menstrual period date could be reconstructed and implied a plausible gestation of 17– 52 weeks, last menstrual period was preferred if it agreed with the gestational-age-implied start within seven days, or if the latter was unavailable; otherwise gestational start was set to the birth date minus the recorded gestational weeks.

Study entry was defined by gestational start. Gestational start was required to occur on or after 1 January 1983. At the close of the study period, eligibility was determined prospectively: gestational start had to fall early enough that a pregnancy continuing to 41 weeks and 6 days — a maximum potential follow-up of 293 days — could still be observed before the natality file ends on 31 December 1988. This yields a latest eligible gestational start of 13 March 1988. This approach is superior to a delivery-year cohort definition, of which a pregnancy conceived late in the observation window enters the data only if it delivers early while a full-term counterpart conceived on the same day delivers after the file ends and is never observed. Mean first trimester exposure is also superior to a rolling average across gestation or the largest nitrate measure observed during gestation, purely because longer gestation (an outcome of interest) will have more time to be exposed to more nitrate measures, increasing the likelihood of exposure to higher nitrate levels; ultimately creating systematic confounding between timing, exposure, and outcomes. This approach avoids sample membership and nitrate exposure becoming a function of gestational duration, and uses a fixed window of exposure in the first trimester to sever that dependence and mitigate bias common to studies leveraging the full gestational period.

### Birth Record Sample Inclusion Criteria

The linked birth cohort was restricted to records with a valid birth date, maternal residence that mapped to a county, a valid gestational start date and gestational age between 20-41 weeks, and a first trimester window falling entirely inside the water panel with at least one observed county-quarter nitrate measure. Births were further restricted to singleton plurality with a gestational age between 20-41 weeks. Pregnancies with any observed nitrate measure >= 10 mg/L or whose completed mean reached or exceeded 10 mg/L in the first trimester were excluded. Two other inclusion criteria were implemented to overcome issues with data sparseness and potential confounding from unmeasured health or socioeconomic factors. Only births to White mothers were included. Other racial/ethnic subgroups in 1980s Iowa are too small (3%) and too geographically concentrated to support county-by-year fixed effects. Only pregnancies with prenatal care initiated by month 5 were retained, fixing care timing across the comparison and removing a channel through which exposure and outcome could be jointly related to care-seeking. Finally, pregnancies with missing birthweight and control variable information were excluded.

### Exposure assignment

The first trimester presents a period when proposed nitrate mechanisms (oxidative stress and methemoglobin-mediated oxygen reduction) may influence birth outcomes[6,10,31,32]. In this study, each pregnancy’s first trimester window, gestational days 0 through 97, was linked to the county-quarter panel. Because a 98-day window spans 2-3 calendar quarters, exposure was assigned based on the later of first trimester start and quarter start and ended on the earlier of first trimester end and quarter end. Then, the inclusive number of overlap days was summed. Exposure was defined as the overlap-day-weighted mean of nitrate measures. Following existing evidence, nitrate measures were modelled as categories: ≤0.1 mg/L (reference, lowest estimated exposure category), greater than 0.1 and below 5 mg/L, and 5 mg/L or above (and, by the analytic restriction, below 10 mg/L)[10,11].

### Statistical Analysis

Four binary outcomes were low birthweight (<2,500 g), very low birthweight (<1,500 g), preterm birth (<37 weeks), and very preterm birth (<32 weeks). Each outcome was estimated by linear probability model with high-dimensional fixed effects county-birth year, which permits each county’s baseline risk to move freely from year to year, and birth year-conception quarter, which absorbs statewide seasonality in conception timing interacted with year. Models adjust for birth parity (first birth, second birth, third or higher birth), maternal education (no high school degree, high school degree only, more than a high school degree), infant sex, marital status, and maternal age. Robust standard errors were clustered at the county level. Average marginal effects were estimated for each exposure category relative to the reference. Because the analyses involved two exposure tests, family-wise adjusted p-values and confidence intervals were computed using the Šidák method, a Bonferroni-type correction that controls the family-wise error rate across the two comparisons[27]. Both uncorrected and Šidák corrected estimates are reported.

### Sensitivity Checks

#### Binary Nitrate Exposure

As a sensitivity analysis, the fixed-effects models were re-estimated using a binary nitrate exposure specification comparing compliant first-trimester concentrations ≥5 mg/L with concentrations <5 mg/L. The same model specification was retained for each exposure definition and for all four outcomes: preterm birth, very preterm birth, low birthweight, and very low birthweight. For inference, permutation tests were conducted within each county with 9,999 replications.

#### Romano-Wolf Permutation Inference Test

Standard approaches to correcting for multiple tests assume independence of outcomes[33]. The four birth outcomes in this study were measured in the same pregnancies and are inherently dependent. For example, very preterm birth is nested within preterm birth. So, the issue of multiple tests with these binary exposure comparisons was addressed using randomization-based inference, specifically the Romano-Wolf permutation test[34]. This Romano-Wolf stepdown algorithm accounts for joint dependence of the test statistics, while providing balance between the family-wise error rate and statistical power with correlated outcomes[35]. Preserving joint dependence of the binary exposure associations, this sensitivity check controlled the family-wise error rate across the four outcome-specific tests using the Romano–Wolf stepdown procedure implemented in Stata[36]. Adjusted p-values were calculated from studentized bootstrap statistics using the stepdown maximum-statistic procedure, with 9,999 bootstrap replications.

#### Completed and Observed Nitrate Measures by Observed Nitrate Days

A second sensitivity check tested robustness between completed and observed nitrate measures. Here, the study re-estimates the binary exposure models using directly observed nitrate measurements and completed (primary) measurements, then by restricting to pregnancies with varying levels of first-trimester observation coverage ^0, ≥30, ≥60, or ≥90 days observed). In these sensitivity checks, exposure was specified as a binary indicator for a first-trimester mean at or above 5 mg/L, so that the observed and completed measures are directly comparable and the contrast does not depend on the size of the non-detect reference group. Specifications, covariates, and fixed effects were otherwise unchanged. Because these robustness checks aimed to assess the comparability between completed and observed nitrate exposure measures, and were not used as primary tests, the estimates are reported without correction for multiple comparisons.

## Results

### Summary Statistics

The final analytic sample included 113,328 births (Figure 1). The mean first trimester nitrate exposure was 2.806 mg/L (SD 2.388, range 0 to <10.0). The distribution was right-skewed (skewness 0.69) with substantial mass near zero: the 5th percentile was 0 mg/L, the 10th percentile 0.13, the median 2.21, the 75th percentile 4.44, the 90th percentile 6.53, and the 99th percentile 8.44 mg/L. By exposure category, 10,423 pregnancies (9.2%) fell in the reference group at or below 0.1 mg/L, 80,038 (70.6%) in the intermediate group above 0.1 and below 5 mg/L, and 22,867 (20.2%) at or above 5 mg/L. Mean birthweight was 3,447.0 g and mean gestational age 39.0 completed weeks. Low birthweight occurred in 4.3% of births, very low birthweight in 0.6%, preterm birth in 7.7%, and very preterm birth in 1.1%. See table 1 for full summary statistics.

**Figure 1.**
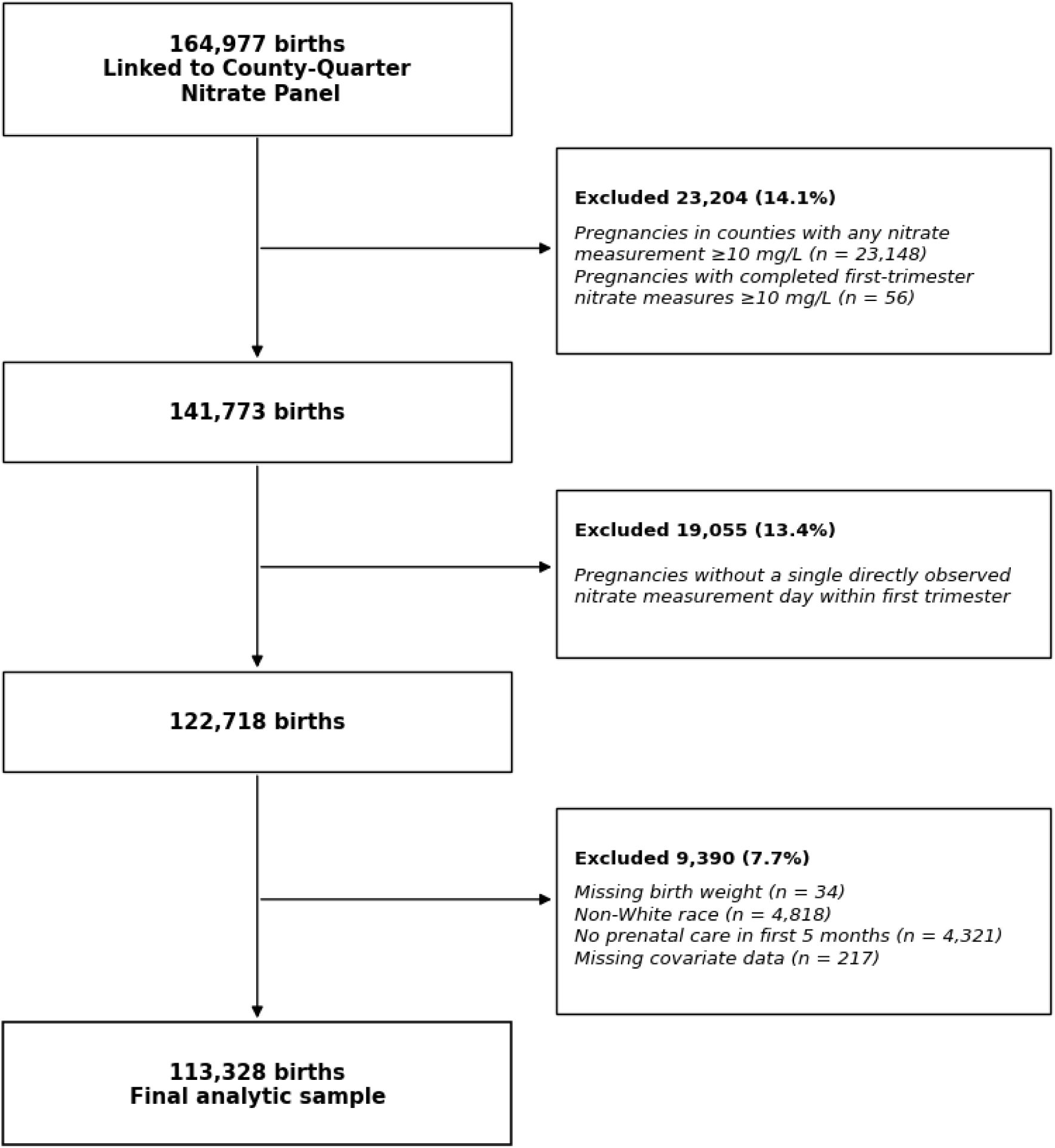
Analytic sample inclusion criteria flow chart. The initial sample included 164,977 births linked to the county-quarter nitrate exposure panel. The linked first trimester window covers 1983-Q1 to 1988-Q2. Final sample included births from 96 of 99 Iowa counties. Cass, Lyon, and Woodbury counties dropped from the sample during the inclusion process. Excluded 23,204 births with observed nitrate concentrations >0 0 mg/L, including 23,148 pregnancies with any observed nitrate measurement >0 m mg/L and 56 additional pregnancies with completed first-trimester nitrate measures >0 0 mg/L. Then excluded 19,055 pregnancies without any directly observed nitrate measurement day during the first trimester. Finally, 9,390 births were excluded because of missing birthweight (n = 34), non-White maternal race (n = 4,818), no prenatal care during the first 5 months of pregnancy (n = 4,321), or missing covariate data (n = 217), yielding a final analytic sample of 113,328 births.

**Table 1.** Characteristics of the analytic sample (N = 113,328)

| Variable | N | Mean |
| --- | --- | --- |
| First trimester nitrate, mg/L |  | 2.806 |
| ≤0.1 mg/L (reference) | 10,423 | 9.20% |
| >0.1 to <5 mg/L | 80,038 | 70.63% |
| ≥5 mg/L | 22,867 | 20.18% |
| Birthweight, g |  | 3,447.0 |
| Gestational age, weeks |  | 39.05 |
| Low birthweight (<2,500 g) | 4,878 | 4.30% |
| Very low birthweight (<1,500 g) | 724 | 0.64% |
| Preterm birth (<37 weeks) | 8,726 | 7.70% |
| Very preterm birth (<32 weeks) | 1,289 | 1.14% |
| Maternal age, years |  | 26.40 |
| Second live birth | 40,608 | 35.83% |
| Third or higher live birth | 29,130 | 25.70% |
| Maternal education, 12 years | 49,701 | 43.86% |
| Maternal education, >12 years | 52,256 | 46.11% |
| Male infant | 58,179 | 51.34% |
| Married | 100,260 | 88.47% |

*Note: Continuous variables report the sample mean; binary and categorical variables report the number and percentage of births in each category. All variables are measured for the 113,328 births in the analytic sample. First-trimester nitrate is the overlap-day-weighted mean completed county-quarter concentration across gestational days 0–97.*

### Primary Results

Estimates from the fixed-effects linear probability models are reported in Table 2 as average marginal effects in percentage points, relative to the reference category of 0.1 mg/L or below. The estimation sample comprised 113,328 births across 96 counties. Figures 2-3 visualize the estimated associations.

**Figure 2.**
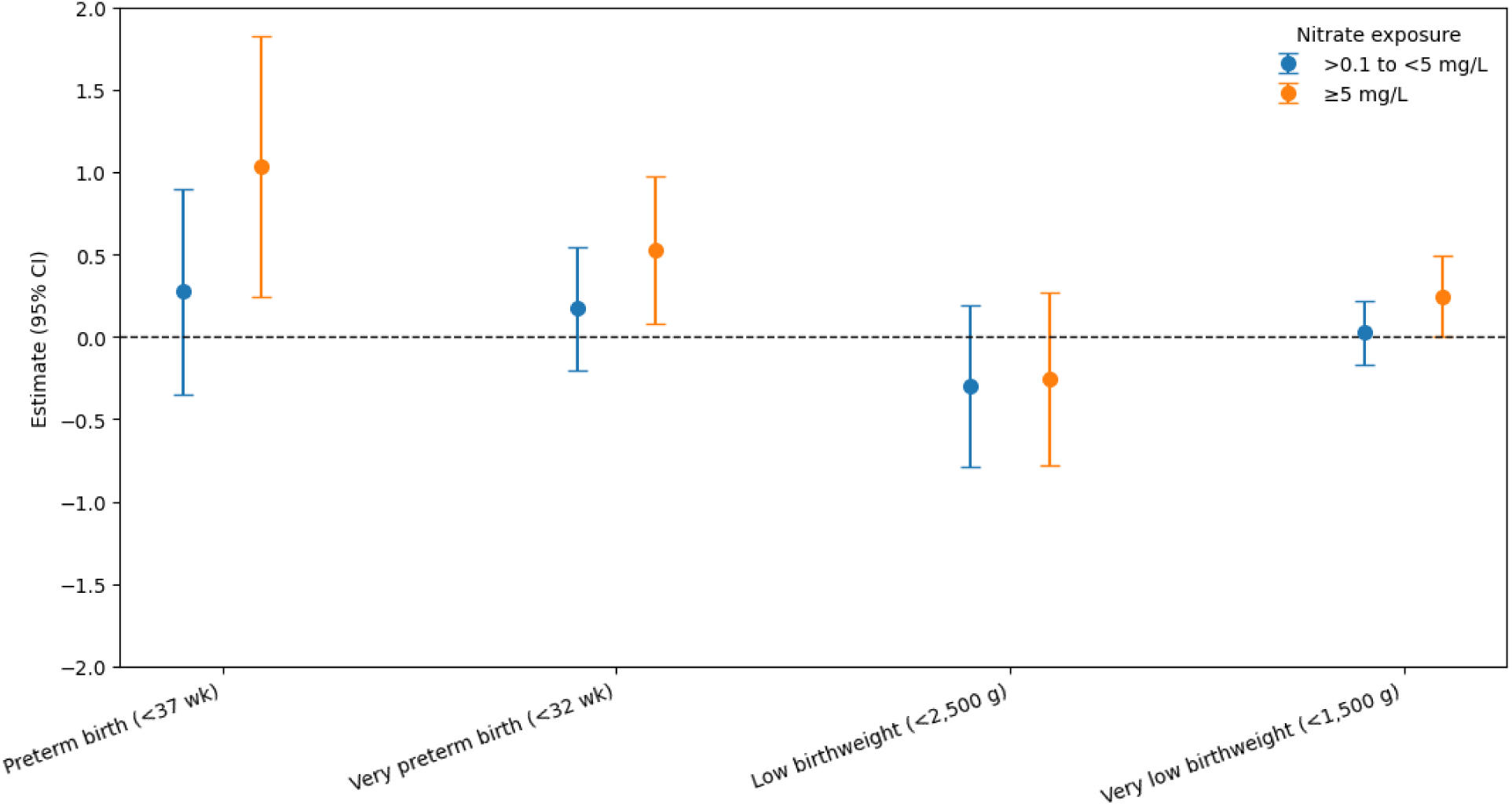
Associations between nitrate exposure and adverse birth outcomes. Points represent estimated percentage-point differences in the probability of each birth outcome relative to nitrate exposure 00.1 mg/L; error bars represent unadjusted 95% confidence intervals. Estimates are shown separately for nitrate concentrations >0.1 to <5 mg/L and >5 mg/L.

**Figure 3.**
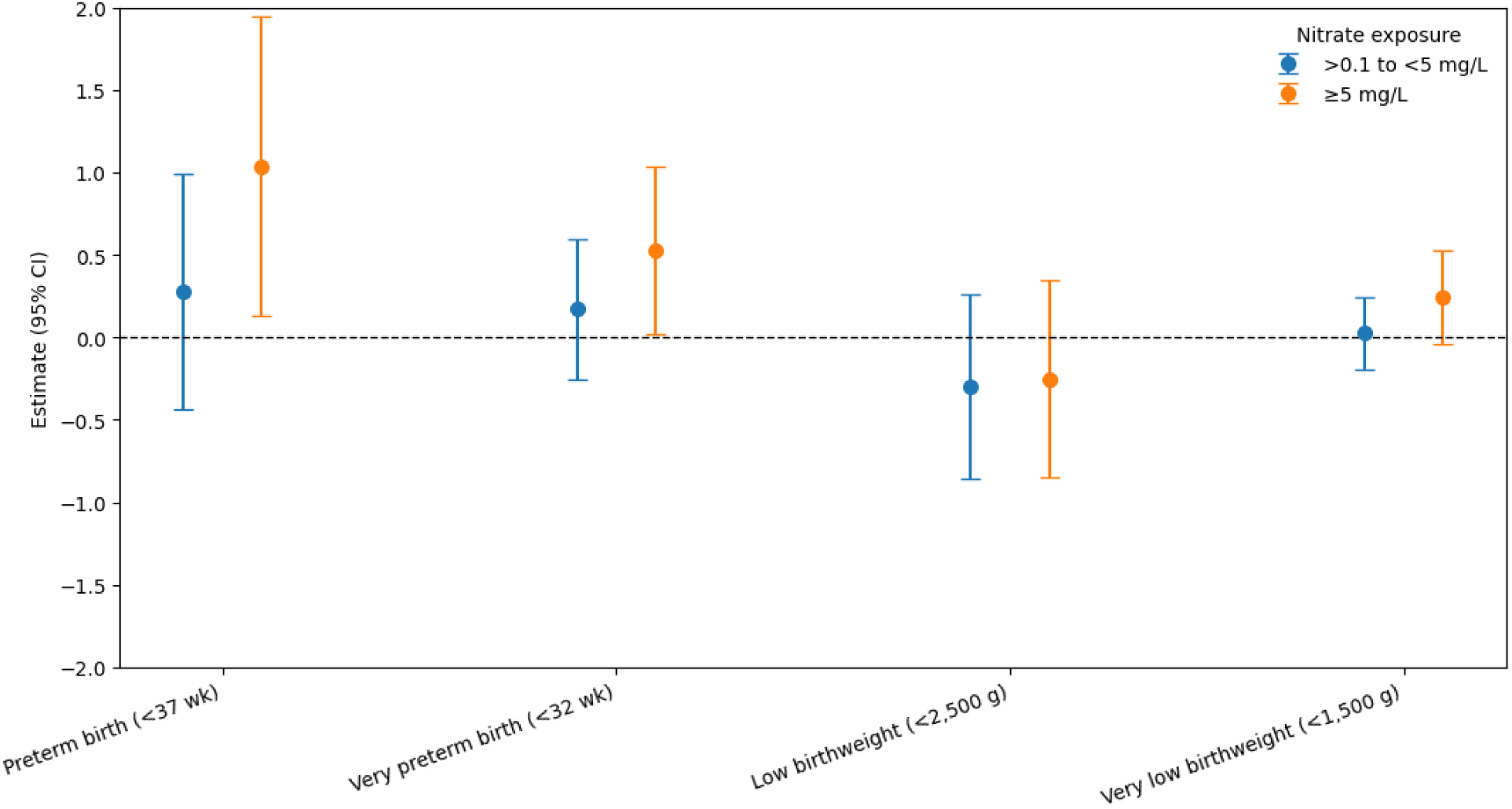
Associations between nitrate exposure and adverse birth outcomes with Šidák-adjusted confidence intervals. Points represent estimated percentage-point differences in the probability of each birth outcome relative to nitrate exposure 00.1 mg/L; error bars represent Sidak-adjusted 95% confidence intervals. Estimates are shown separately for nitrate concentrations >0.1 to <5 mg/L and >5 mg/L.

**Table 2.**
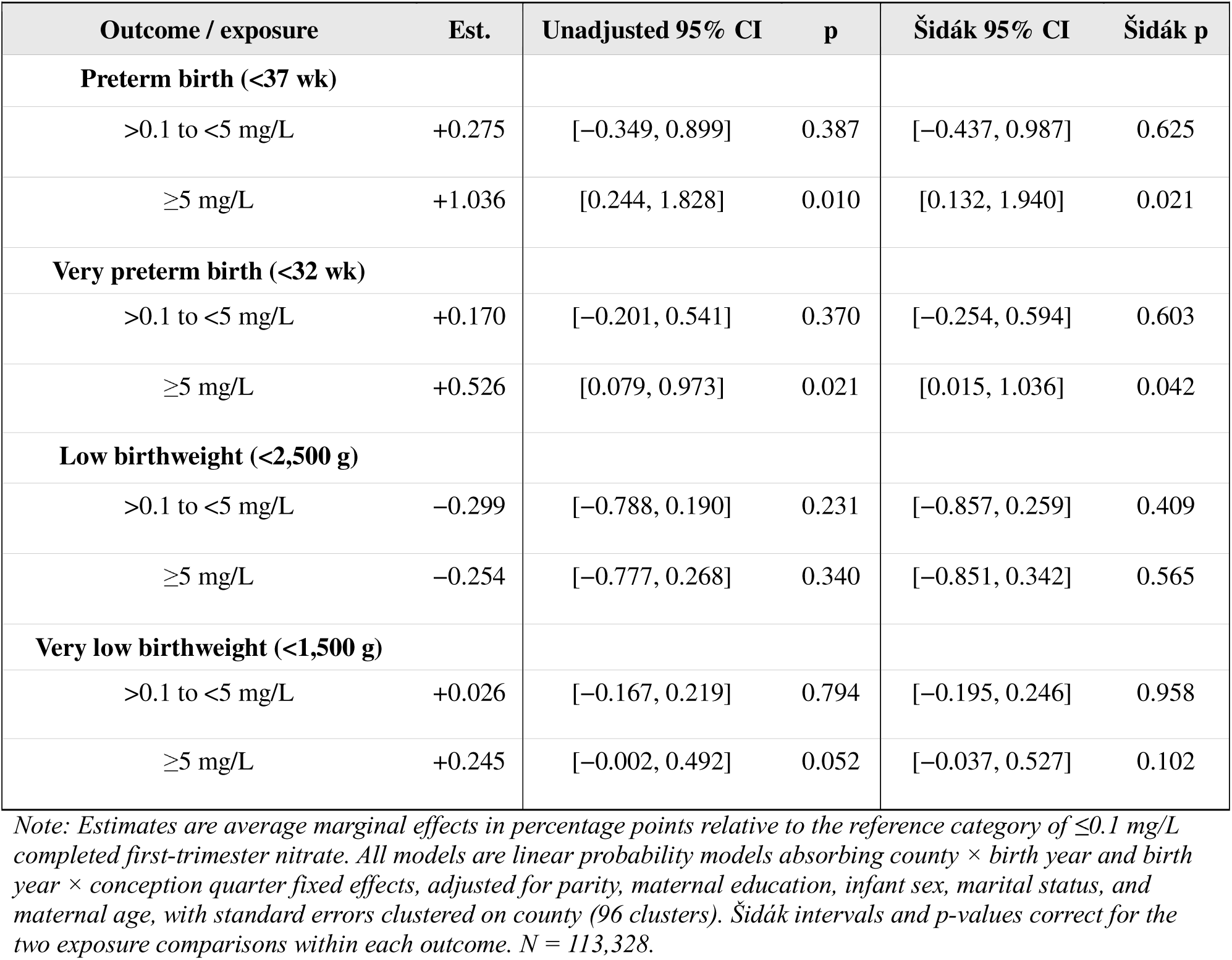
Average marginal effects of first-trimester nitrate exposure on birth outcomes.

#### Preterm birth

First-trimester nitrate at or above 5 mg/L was associated with a 1.036 percentage-point increase in the probability of preterm birth (95% CI 0.244 to 1.828; p = 0.010). This represents a 13.5% relative increase from the mean. The estimate remained statistically significant after correction for multiple comparisons (Šidák-adjusted 95% CI 0.132 to 1.940; adjusted p = 0.021). The intermediate category, above 0.1 and below 5 mg/L, was not associated with preterm birth (0.275 percentage points; 95% CI −0.349 to 0.899; p = 0.387; adjusted 95% CI −0.437 to 0.987; adjusted p = 0.625). The two exposure category associations differed significantly from each other (F(1,95) = 6.39; p = 0.0131).

#### Very preterm birth

Exposure at or above 5 mg/L was associated with a 0.526 percentage-point increase in the probability of very preterm birth (95% CI 0.079 to 0.973; p = 0.021). This represents a 46.2% relative increase from the mean. The estimate remained statistically significant after correction for multiple comparisons (Šidák-adjusted 95% CI 0.015 to 1.036; adjusted p = 0.042). The intermediate category showed no association (0.170 percentage points; 95% CI −0.201 to 0.541; p = 0.370; adjusted 95% CI −0.254 to 0.594; adjusted p = 0.603). The two exposure category associations differed significantly from each other (F(1,95) = 8.69; p = 0.0040).

#### Low birthweight

Neither exposure category was associated with low birthweight and the two exposures did not differ from each other (F(1,95) = 0.09; p = 0.7671). The estimate for exposure at or above 5 mg/L was −0.254 percentage points (95% CI −0.777 to 0.268; p = 0.340; adjusted 95% CI −0.851 to 0.342; adjusted p = 0.565), and for the intermediate category −0.299 percentage points (95% CI −0.788 to 0.190; p = 0.231; adjusted 95% CI −0.857 to 0.259; adjusted p = 0.409).

#### Very low birthweight

Exposure at or above 5 mg/L was associated with a 0.245 percentage-point increase in the probability of very low birthweight, of similar relative magnitude to the very preterm birth estimate but not statistically distinguishable from zero (95% CI −0.002 to 0.492; p = 0.052; adjusted 95% CI −0.037 to 0.527; adjusted p = 0.102). The intermediate category showed no association (0.026 percentage points; 95% CI −0.167 to 0.219; p = 0.794; adjusted 95% CI −0.195 to 0.246; adjusted p = 0.958). The two exposure category associations differed significantly from each other (F(1,95) = 7.47; p = 0.0075).

### Sensitivity Checks

Compared to reference category <5 mg/L, exposure to nitrate ≥5 mg/L was associated with a 0.76 [CI = 0.17 to 1.35] percentage-point higher probability of preterm birth, a 0.35 [CI = 0.12 to 0.59] percentage-point higher probability of very preterm birth, and a 0.22 [CI = 0.06 to 0.38] percentage-point higher probability of very low birthweight. There was no association between exposure to nitrate ≥5 mg/L and the probability of low birthweight. Table 3 reports these estimated associations, along with inferential p-values from the stand alone regression, permutation tests, and the Romano-Wolf adjustment.

**Table 3.**
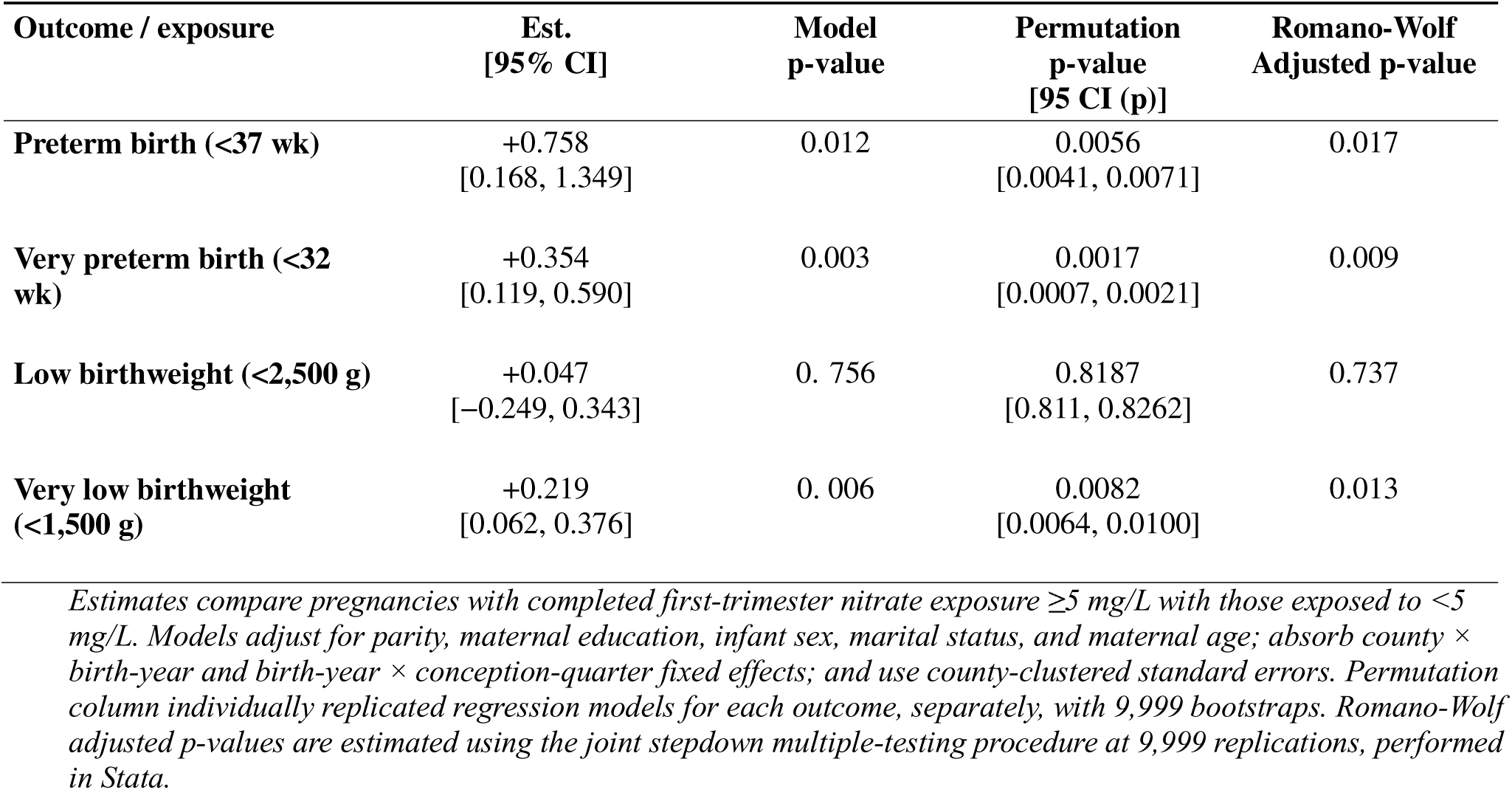
Sensitivity analysis of first-trimester nitrate exposure 55 mg/L and adverse birth outcomes (Romano-Wolf Permutation)

In the preferred 9,999 Wolf adjustment, the inference for the preterm birth (p = 0.017), very preterm birth (p = 0.009), and very low birth weight (p = 0.013) remain statistically significant at p < 0.05. In contrast, there was no evidence of an association with low birthweight (model p=0.756; Romano-Wolf-adjusted p=0.728). Thus, the associations with preterm birth, very preterm birth, and very low birthweight remained below the 0.05 family-wise significance threshold after dependence-aware correction for the four outcomes, whereas the low-birthweight association remained null. Even if a Sidak correction were applied post-hoc (alpha = 0.0127), again not recommended given the dependence of outcomes, the very preterm birth estimate remains statistically significant. Taken together, the primary analysis and sensitivity checks suggest that the statistical significance of these estimates are not due to random chance driven by multiple hypotheses.

Table 4 reports the robustness check comparing completed and observed nitrate measures. Requiring at least 30, 60, or 90 of the 98 first-trimester days to fall within a directly observed county-quarter reduced the sample from 113,328 to 103,956, 93,654, and 83,688 births, respectively. Across all four coverage thresholds and both exposure measures, the two gestational-duration associations were stable in sign, magnitude, and statistical significance. For preterm birth, the estimate for exposure at or above 5 mg/L ranged from 0.667 to 0.814 percentage points and remained statistically significant in all eight specifications (p ≤ 0.016). For very preterm birth, the estimate ranged from 0.296 to 0.354 percentage points and likewise remained significant throughout (p ≤ 0.014). Estimates constructed from directly observed county-quarters alone were nearly indistinguishable from those using the completed panel at every threshold, differing by no more than 0.06 percentage points for preterm birth and 0.05 percentage points for very preterm birth. Low birthweight was null in all eight specifications. Very low birthweight was positive throughout and statistically significant in six of eight specifications; the two exceptions fell at the boundary of conventional significance, using the observed-only measure at the 30-day and 60-day thresholds (p = 0.052 and p = 0.051). That estimates constructed without any completed input reproduce those from the completed panel, and that both are insensitive to progressively demanding observation requirements, indicates that the reported associations are not artifacts of the spatiotemporal imputation (Figure 4).

**Figure 4.**
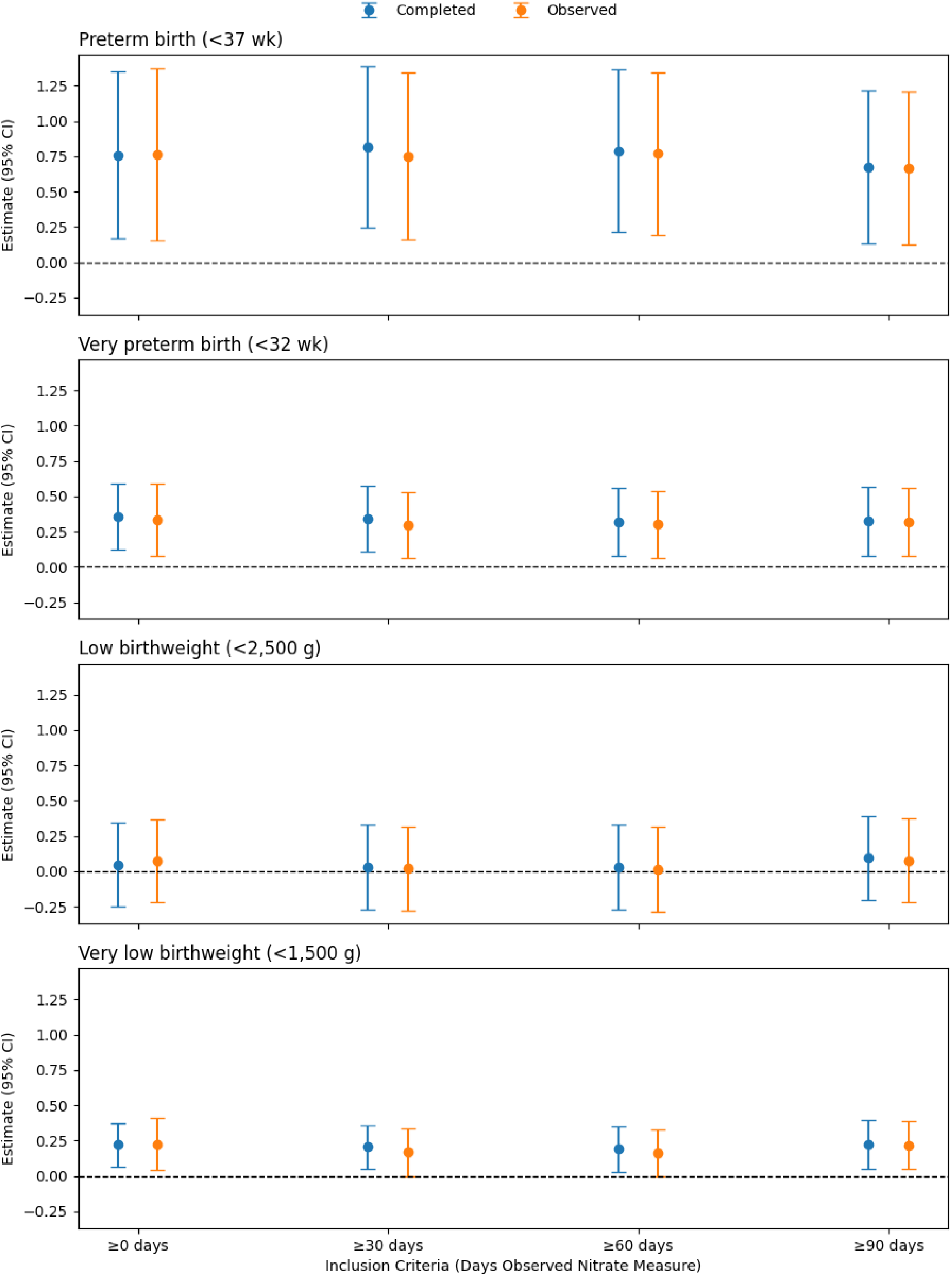
Comparing associations between completed and observed nitrate measures, by coverage days of observed nitrate measures in first trimester. Points represent average marginal effects in percentage points for first-trimester mean nitrate concentrations 55 mg/L relative to <5 mg/L; error bars represent 95% confidence intervals. “Completed” uses the completed county-quarter panel, including spatiotemporally completed values, whereas “Observed” uses directly observed county-quarters only. The x-axis indicates the minimum number of first-trimester days (of 98) falling within directly observed county-quarters.

**Table 4.**
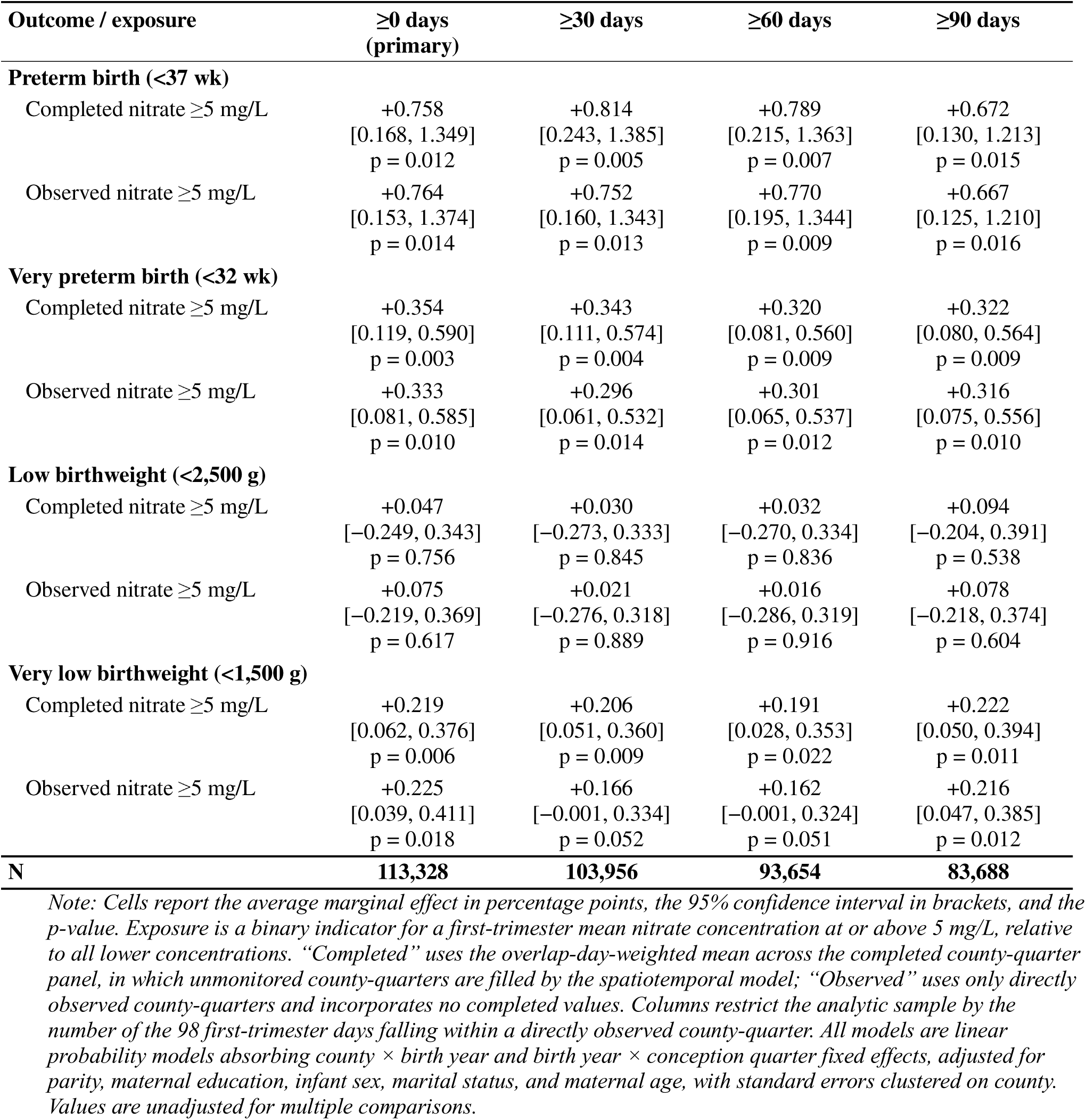
Average marginal effects of first-trimester nitrate exposure at or above 5 mg/L, by exposure measure and first-trimester observation coverage.

| Outcome / exposure | ≥0 days<br>(primary) | ≥30 days | ≥60 days | ≥90 days |
| --- | --- | --- | --- | --- |
| **Preterm birth (<37 wk)** |  |  |  |  |
| Completed nitrate ≥5 mg/L | +0.758<br>[0.168, 1.349]<br>p = 0.012 | +0.814<br>[0.243, 1.385]<br>p = 0.005 | +0.789<br>[0.215, 1.363]<br>p = 0.007 | +0.672<br>[0.130, 1.213]<br>p = 0.015 |
| Observed nitrate ≥5 mg/L | +0.764<br>[0.153, 1.374]<br>p = 0.014 | +0.752<br>[0.160, 1.343]<br>p = 0.013 | +0.770<br>[0.195, 1.344]<br>p = 0.009 | +0.667<br>[0.125, 1.210]<br>p = 0.016 |
| **Very preterm birth (<32 wk)** |  |  |  |  |
| Completed nitrate ≥5 mg/L | +0.354<br>[0.119, 0.590]<br>p = 0.003 | +0.343<br>[0.111, 0.574]<br>p = 0.004 | +0.320<br>[0.081, 0.560]<br>p = 0.009 | +0.322<br>[0.080, 0.564]<br>p = 0.009 |
| Observed nitrate ≥5 mg/L | +0.333<br>[0.081, 0.585]<br>p = 0.010 | +0.296<br>[0.061, 0.532]<br>p = 0.014 | +0.301<br>[0.065, 0.537]<br>p = 0.012 | +0.316<br>[0.075, 0.556]<br>p = 0.010 |
| **Low birthweight (<2,500 g)** |  |  |  |  |
| Completed nitrate ≥5 mg/L | +0.047<br>[-0.249, 0.343]<br>p = 0.756 | +0.030<br>[-0.273, 0.333]<br>p = 0.845 | +0.032<br>[-0.270, 0.334]<br>p = 0.836 | +0.094<br>[-0.204, 0.391]<br>p = 0.538 |
| Observed nitrate ≥5 mg/L | +0.075<br>[-0.219, 0.369]<br>p = 0.617 | +0.021<br>[-0.276, 0.318]<br>p = 0.889 | +0.016<br>[-0.286, 0.319]<br>p = 0.916 | +0.078<br>[-0.218, 0.374]<br>p = 0.604 |
| **Very low birthweight (<1,500 g)** |  |  |  |  |
| Completed nitrate ≥5 mg/L | +0.219<br>[0.062, 0.376]<br>p = 0.006 | +0.206<br>[0.051, 0.360]<br>p = 0.009 | +0.191<br>[0.028, 0.353]<br>p = 0.022 | +0.222<br>[0.050, 0.394]<br>p = 0.011 |
| Observed nitrate ≥5 mg/L | +0.225<br>[0.039, 0.411]<br>p = 0.018 | +0.166<br>[-0.001, 0.334]<br>p = 0.052 | +0.162<br>[-0.001, 0.324]<br>p = 0.051 | +0.216<br>[0.047, 0.385]<br>p = 0.012 |
| **N** | **113,328** | **103,956** | **93,654** | **83,688** |

*Note: Cells report the average marginal effect in percentage points, the 95% confidence interval in brackets, and the p-value. Exposure is a binary indicator for a first-trimester mean nitrate concentration at or above 5 mg/L, relative to all lower concentrations. “Completed” uses the overlap-day-weighted mean across the completed county-quarter panel, in which unmonitored county-quarters are filled by the spatiotemporal model; “Observed” uses only directly observed county-quarters and incorporates no completed values. Columns restrict the analytic sample by the number of the 98 first-trimester days falling within a directly observed county-quarter. All models are linear probability models absorbing county × birth year and birth year × conception quarter fixed effects, adjusted for parity, maternal education, infant sex, marital status, and maternal age, with standard errors clustered on county. Values are unadjusted for multiple comparisons.*

The estimates in Table 4 are not simply more precise versions of those in Table 2; they answer a slightly different question. The primary models contrast exposure at or above 5 mg/L against non-detect pregnancies alone (<0.1 mg/L; 10,423 births), whereas the binary models contrast exposure at or above 5 mg/L against all pregnancies below 5 mg/L (90,461 births). For very low birthweight, the estimate is nearly unchanged while the comparison group is almost nine times larger, and the association reaches conventional significance in Table 4. For low birthweight the estimate changes signs between the two tables while remaining null in both.

## Discussion

Among 113,328 singleton births to White Iowa mothers delivered between 1983 and 1988, first-trimester exposure to compliant drinking-water nitrate at or above 5 mg/L was associated with increased probability of preterm birth compared to lowest exposure category, a 13.5% relative increase that persisted after correction for multiple comparisons. The same exposure category was associated with increased very preterm birth, a 46.2% relative increase which also remained statistically significant after correction for multiple comparisons. Exposure above 0.1 mg/L but below 5 mg/L was not associated with any outcome. No association was detected for low birthweight. All significant associations reflect exposure to compliant nitrate under current regulations.

The sensitivity analyses address the concern that modelled county-quarter values, rather than directly observed exposure, generate the reported associations. Estimates built without any modelled input were nearly indistinguishable from those using the completed panel: differing by no more than 0.06 percentage points for preterm birth and 0.05 for very preterm birth. Both were stable as the observation days restrictions tightened.

Roughly 85% of Iowa’s land area is in farmland, cultivated overwhelmingly in corn and soybean and combined with dense livestock production, and a large share of public water systems draw on shallow alluvial and bedrock aquifers vulnerable to agricultural nitrate[16,37,38]. The historical period in this study carries a further methodological advantage. Although nitrate violations have been reported since 1978, requirements for public water systems to monitor and report violations changed in 1993[39]. Measured nitrates in public drinking water in the late 1980s may correspond to concentrations pregnant women drank. Thus, this study presents important, population-based evidence on the association between early prenatal exposure to compliant nitrate on birth outcomes in an intensive agriculture state.

### Comparison with existing evidence

The preterm birth estimates align closely with the evidence from California. Montoya (2025) reported a 1.2 percentage-point increase in preterm birth, a 15% relative increase, from second-trimester exposure to nitrate between 5 and 10 mg/L in California community water systems; the present estimate is 1.0 percentage points and 13.5%[11]. This convergence is notable. The estimates were derived from different states with different water-system infrastructure, in different decades. That two independent bodies of data separated by decades and half a continent produce sub-limit preterm effects of nearly identical magnitude strengthens the case that the association is not an artifact of either setting or methodology. It also extends the finding to a high-agriculture context, where nitrate is chronic and seasonally driven rather than episodic, which was the principal gap motivating this current study.

The severity gradient observed here also parallels Sherris et al. (2021). In that within-mother analysis, exposure of 5 to <10 mg/L was associated with a large increase in the odds of spontaneous preterm birth at 20–31 weeks but only a slight increase at 32–36 weeks, an effect concentrated at the severe end of the gestational-duration distribution[10]. The present results show the same ordering in relative terms for preterm birth and very preterm birth outcomes.

Still, differences from the California literature merit attention. Montoya’s measure is an indicator for any single month reaching 5–10 mg/L within a trimester, while the measure here is a 98-day mean at or above 5 mg/L, which requires more sustained measured nitrate elevation throughout the first trimester. Additionally, quarterly monitoring cannot isolate a trimester as precisely as monthly data. Also, quarterly values assigned to gestational days 0–97 describe ambient conditions in the adjacent weeks, and because agricultural nitrate is strongly correlated within a year, a first-trimester measure in these data carries information about concentrations in the surrounding period[40].

### Future research

As research continues to mount that prenatal exposure to compliant nitrate may harm birth outcomes, important questions remain unanswered. Whether early-pregnancy exposure, mid-pregnancy exposure, or sustained exposure across both drives potential harms remains a testable question, one which the present quarterly data can pose but not adequately resolve. The concentration of effects at the severe end of both outcome distributions also invites analyses powered specifically for very preterm birth and very low birthweight, possibly in subpopulations at greater risk of elevated exposure and high risk of adverse birth outcomes. Such research, especially in relatively homogenous agricultural states would require either a longer panel or pooling across states. In that regard, with available data, the existing studies provide methodological guidance for replication across other states which lack substantial evidence using population-based data and causal inference methods. More specific to this current study, contemporary replication would improve the policy relevance of this research. Although the 1980s offer unusually clean correspondence between measured and consumed concentrations, quantifying how much modern treatment and source-switching attenuate that correspondence would clarify how best to prioritize strategies for reducing the potential harm from nitrates on pregnant women today.

### Policy and practice implications

The findings reinforce the argument that the 10 mg/L standard may not protect against the developmental effects of prenatal nitrate exposure[6,9,41,42]. The association reported here is identified entirely below the current threshold, among pregnancies whose first-trimester exposure never reached the regulatory limit. Montoya (2025) drew the explicit implication that a limit below 5 mg/L would prevent nitrate-attributable adverse birth outcomes[11]. The present results are consistent with that argument and extend its evidentiary base to a chronic, agriculturally driven region. Because the standard was derived from acute infant methemoglobinemia rather than from fetal development, and has stood unrevised, a reassessment that takes developmental endpoints as the basis for the limit appears warranted. At minimum, the bulk of research should motivate policymakers and regulators to at least officially include pregnant women as a population at risk of harm from consuming elevated nitrates in drinking water.

Monitoring and public notification are currently organized around detecting exceedances of 10 mg/L, so systems operating persistently between 5 and 10 mg/L generate no urgent violation notice. Reporting sub-limit concentrations to the public, and to prenatal care providers, could give pregnant women and clinicians information that the current framework lacks. Although consumer reports already provide information about nitrate concentrations above 5 mg/L, further research could inform whether prenatal-risk communication should be incorporated into existing reporting practices. Relatedly, whether exposure in this range is modifiable at low cost through targeted use of alternative water sources during early pregnancy could be explored by municipal drinking water administrators and local officials. Because nitrate in agricultural regions is strongly seasonal, and because the exposures identified here are sustained rather than transient, source-water protection and nutrient-management measures that reduce baseline concentrations in vulnerable aquifers would be expected to shift a substantial share of the exposure distribution below the range associated with harm. Finally, from a public health perspective, we as a field can improve how we communicate risks of nitrate. Current public health strategies center around compliance, so the absence of a violation could be interpreted by the public as an absence of risk. Yet, the associations reported here, and in both California studies, are identified entirely among systems that were fully compliant. Annual water-quality reports compound the problem by describing the prior year’s concentrations months after the fact, which is of limited use to someone deciding what to drink during a pregnancy already underway. The other extreme, rapid communication of any nitrate measure without context could be equally unhelpful. And because the first trimester often precedes the initiation of prenatal care, and sometimes the recognition of pregnancy itself, information delivered at a first prenatal visit arrives after a possibly critical window for reducing nitrate exposure. As the public waits for regulatory reform, public health systems communicating sub-limit concentrations in real time, and in terms people can act on, could be implemented as a low-cost complement for pregnant women, now.

### Limitations

Despite improvements from the original study by Semprini (2025), the present study’s results should be interpreted understanding its limitations. Nitrate was assigned at the county-quarter level rather than at the level of the individual water system or household, missing potential sub-county variation in exposure to nitrate. Information on private wells, which serve many households in rural Iowa, was also not included in this data[38,43]. Migration within a county could also not be measured. Because most of these sources of error are plausibly unrelated to birth outcomes conditional on the fixed effects, these sources of error are expected to be non-differential and would ordinarily attenuate estimates toward the null, which suggests the associations reported here are more likely understated than overstated. Quarterly monitoring also limits temporal precision: the data cannot resolve short-lived concentration spikes within a quarter, nor separate the first trimester from adjacent weeks as sharply as monthly data allow. As described in the methods, part of the final birth cohort’s first trimester exposure days were constructed by predictive modeling but the similarity between the completed and observed nitrate exposure estimates provides greater confidence that the primary findings were not purely a result of the modelling procedure. Restricting the analytic sample to births from White mothers with prenatal care initiated by month 5 was decided to homogenize the sample on dimensions strongly associated with baseline obstetric risk and with care-seeking behavior, and minimize sparse data in homogenous counties. The limitation here is generalizability, and the estimates should not be extrapolated to other racial and ethnic groups, to pregnancies with late or absent prenatal care, or to concentrations at or above the regulatory limit. Further, these estimates may not extend beyond Iowa. The present study also lacked clinically relevant variables available on modern birth records, which could have been used to identify and exclude higher risk pregnancies. Residual confounding by factors that move with the agricultural calendar could not be included given the time period. Finally, the study period ends in 1988. The historical setting in Iowa may help with internal validity and population significance, but may also limit how well this evidence generalizes to other states today.

## Conclusions

With existing research showing prenatal exposure to compliant nitrate may be associated with increased risk of adverse birth outcomes, this present study aimed to improve upon prior work and extend the evidence base to a state with some of the highest levels of nitrate in the nation. Among 113,328 singleton births in Iowa between 1983 and 1988, first-trimester exposure to drinking-water nitrate at or above 5 mg/L was associated with increased probability of preterm birth and very preterm birth. These associations persisted after correction for multiple comparisons and were generally consistent with recent research in California. Together, the available evidence raises questions as to whether the current regulatory and reporting approaches adequately protect the health of pregnant women and their babies.

## Data Availability Statement

All data and code necessary to reproduce the analyses are publicly available. The original natality data used to construct the birth records are publicly available through the National Bureau of Economic Research (NBER) Natality Data repository. The original water-quality data was requested from CHEEC and can be found, unaltered from the original in the corresponding author’s data repository. Note, the original water data from CHEEC did not contain a data dictionary and had incorrect geocode labels (i.e., swapped water sample latitude and longitude). The original finished water data in this study includes the correct geocode labels. Both the filtered, cleaned finished water data and the final county-quarter nitrate exposure panel are archived in the Open Science Framework (OSF) repository [https://doi.org/10.17605/OSF.IO/P39W4].

All author-generated code used for data preparation and analysis is publicly available in the GitHub repository [https://github.com/jsemprini/Iowa-Water-Nitrate-Births-8388-Final]. This repository includes Python notebooks used to clean and process the water-quality data, construct the county-quarter nitrate exposure panel, prepare the natality records, and link nitrate exposures to birth records. The Python notebooks are configured for use in Google Collaboratory environments, with source data accessed through Google Drive as described in the repository documentation. The repository also contains the Stata do-file used to conduct the statistical analyses reported in the manuscript.

## Ethics

Not human subjects research.

